# Developing the Longitudinal Study of Aging in Guatemala (ELEGUA): Rationale and pilot protocol

**DOI:** 10.64898/2026.08.26.26361136

**Authors:** Karen Corzantes, Karyn Choy, Sara Adar, Luis Fernando Castellanos, Alden L. Gross, Kenneth M. Langa, Peter Rohloff, Bas Weerman, Emily Briceño, Manuel Ramírez-Zea, Jere Behrman, David Flood

## Abstract

**Introduction:** Guatemala is the most populous country in Central America and a setting with unique opportunities for aging research. Approximately 40% of Guatemala’s population is Indigenous Maya, who together speak 22 Mayan languages. Currently, there is no population-based aging study in Guatemala and few aging studies in Latin America among Indigenous populations. The Longitudinal Study of Aging in Guatemala (ELEGUA) aims to address these gaps by developing a nationally representative, population-based, longitudinal aging study modeled on the Health and Retirement Study and the Harmonized Cognitive Assessment Protocol, adapted to the cultural and linguistic context of Guatemala. The objective of this protocol is to describe the rationale and design of the ELEGUA pilot survey.

**Methods and analysis:** The ELEGUA pilot was a cross-sectional household survey of adults aged 40 years or older in Tecpán, Guatemala. Tecpán was chosen because its diverse population facilitated testing of study procedures in both Spanish and Kaqchikel, a common Mayan language. The survey included up to 600 households sampled using a multistage stratified cluster design. Within each household, one individual aged 40 years or older was selected, with oversampling of adults aged 55 years or older. This respondent completed a comprehensive questionnaire, including detailed cognitive tests, and provided physical measurements and a venous blood sample. Household respondents provided information on household economics and family structure, and an informant reported on the individual respondent’s cognitive function. Data were collected using a computer-assisted personal interviewing system. Planned analyses include survey-weighted descriptive statistics and psychometric evaluation of the cognitive assessments.

**Ethics and dissemination:** Ethics approval was obtained from the ethics committees of the Institute of Nutrition of Central America and Panama, Maya Health Alliance, and the University of Michigan. Results will be disseminated through publications in peer-reviewed journals and presentations to local, national, and international audiences.

**Strengths and limitations of this study:**

- The Longitudinal Study of Aging in Guatemala (ELEGUA) aims to be the first population-based aging study in Guatemala, with instruments adapted from the Health and Retirement Study (HRS) International Network of Studies and the Harmonized Cognitive Assessment Protocol (HCAP).
- The pilot survey, including the full HCAP cognitive battery, was administered in Spanish and Kaqchikel, a commonly spoken Mayan language in Guatemala.
- The survey included a rigorous probabilistic multistage stratified cluster sampling design based on national census maps and household listings.
- Field procedures, including computer-assisted personal interviewing, physical measurements, and venous blood collection, were piloted under conditions approximating those of the planned national study.
- As a cross-sectional pilot in a single municipality, the study was designed to test procedures rather than to achieve statistical precision, and findings are not intended to generalize to all regions in Guatemala.

## INTRODUCTION

International longitudinal aging studies illuminate how aging processes, including the development of chronic diseases and dementia, are shaped by the complex interplay of biological, environmental, and social determinants. Many of the most influential of these studies are modeled on the U.S. Health and Retirement Study (HRS) and together form a global research network, the HRS International Network of Studies.^1^ Studies in this network use comparable methods to collect data on the health and well-being of aging adults in more than 40 countries.^2,3^ A growing number of these studies also field a Harmonized Cognitive Assessment Protocol (HCAP), a substudy that includes a detailed neuropsychological battery of tests alongside a cognitive-focused informant interview.^4^ The HCAP battery was designed to be harmonizable across countries to facilitate cross-national research on cognitive health, including comparisons of dementia prevalence, incidence, and risk factors.^4,5^

In Latin America, the Mexican Health and Aging Study^6,7^ and the Costa Rican Longevity and Healthy Aging Study^8^ exemplify the contributions of the HRS International Network of Studies, including HCAP administration. These cohorts have yielded insights into a broad range of scientific questions such as the Hispanic paradox among Mexican immigrants,^9^ exceptional longevity in Costa Rica,^10^ and dementia risk factors in Mexico.^11^ Yet between these two countries lies a gap: there has never been a population-based aging study in Guatemala, the most populous country in Central America and a setting with unique opportunities for aging research. The Longitudinal Study of Aging in Guatemala (ELEGUA, from the Spanish “Estudio Longitudinal de Envejecimiento en GUAtemala”) aims to address this gap.

### Rationale for a longitudinal aging study in Guatemala

Guatemala is a middle-income country of 19 million people at an early but rapidly accelerating stage of the demographic transition.^12^ The country is a compelling setting for a longitudinal aging study as it provides distinctive populations, exposures, and scientific opportunities for aging research.

First, Guatemala fills an important gap in global aging research coverage. The HRS International Network of Studies has broad worldwide reach, yet it currently includes fewer than 20 of the approximately 130 low- and middle-income countries.^13–15^ Within Latin America and the Caribbean, HRS or HCAP studies are established in Brazil,^16^ Mexico,^6^ Costa Rica,^8^ Cuba,^17^ Chile,^18^ the Dominican Republic,^17^ and Puerto Rico^17,19^—all countries or territories wealthier than Guatemala. Foundational evidence on aging and dementia in the region also has come from cross-sectional surveys and community-level cohorts, notably the 10/66 Dementia Research Group studies.^20^ There is a need for new aging studies in Latin America that capture the region’s immense diversity both within and across countries.^21,22^ To date, no longitudinal aging study has been conducted in Guatemala, the most populous country in Central America and one of the poorest countries in Latin America with more than half the population living in poverty.^23^

Second, approximately 40% of Guatemala’s population is Indigenous Maya, among the highest proportions of Indigenous people of any country in the Americas.^24,25^ (Following standard usage,^26^ “Maya” refers to the *people* and “Mayan” to the *languages*.) Indigenous populations globally are severely underrepresented in aging and dementia research, particularly outside high-income Western countries.^27,28^ Indigenous populations have distinctive exposures, biological markers, and genetic heritage, and understanding these factors is important both to promote healthy aging among marginalized Indigenous populations themselves and to advance aging science generally.^29^ For example, Guatemala offers a rare setting in which to characterize how discrimination and trauma, as well as protective cultural factors, shape health and well-being in later life. Maya adults in Guatemala may be at elevated risk of poor aging outcomes because they experience systematic life-course discrimination in education, employment, and health care,^25,30^ and many are now entering later life having lived through the genocide against Maya people during the Guatemalan Civil War (1960-1996).^31^ At the same time, healthy aging may be promoted among Maya adults by physically demanding agricultural and subsistence work that sustains high physical activity into later life,^32^ traditional diets low in ultra-processed foods,^33^ and cultural norms favoring multigenerational households and social connectedness.^34^

A longitudinal aging cohort in Guatemala provides the opportunity to examine how the tapestry of these social, environmental, and genetic factors shapes healthy aging, including cognition, among Indigenous populations.

Third, given the high proportion of Indigenous Maya people, nearly one-third of Guatemalans speak one of the country’s 22 Mayan languages.^24^ Mayan languages differ from Spanish and English in fundamental ways that may affect survey measurement, particularly cognitive assessment. For example, verbs are structured differently from those in Spanish, and counting uses a different numbering system (vigesimal, or base-20)—among many distinctive linguistic features of Mayan languages.^26^ Additionally, few speakers read or write in Mayan, and Mayan-Spanish bilingualism and code-switching are common. To our knowledge, no study in the HRS International Network of Studies administers assessments in an Indigenous language of the Americas. In Mexico, speakers of Indigenous languages who are interviewed in Spanish show worse late-life cognitive performance than monolingual Spanish speakers.^35^ Whether these disparities reflect underlying educational differences, effects of multilingualism, or the measurement properties of cognitive tests developed for Spanish speakers remains unclear. A Guatemalan aging study with assessments in Mayan languages has the potential to disentangle these explanations and advance the science of cognitive measurement in linguistically diverse populations.

Fourth, Guatemalans age with minimal social protections. Guatemala has the lowest pension coverage in the Americas (13% of adults aged 65 years or older),^36^ limited access to health insurance (80% of the population is uninsured),^37,38^ and extreme underinvestment in public health care (government spending on health care, at 2.5% of GDP, is among the lowest levels in Latin America).^39^ Longitudinal data from Guatemala can illuminate the natural history of aging in ways that would be difficult to study in many other contexts.

Fifth, Guatemalan adults are, on average, among the shortest populations in the world.^40^ Adult short stature is a marker of early-life environmental and nutritional deprivation and is causally linked to deleterious adult outcomes including lower educational attainment, lower adult wages, and higher risk of cardiometabolic disease (e.g. obesity, diabetes, and hypertension).^41–43^ Whether these early-life exposures also influence late-life cognition and dementia risk is less established and remains a key question that a longitudinal aging study in Guatemala is well-positioned to investigate.

Finally, Guatemala’s population ages in a context of high international migration and dependence on remittances, which are financial transfers sent by migrants abroad to family members in their home country. Approximately 10% of Guatemalan-born adults reside abroad,^44^ and 6% of households have at least one member living permanently outside the country.^24^ Remittances are equivalent to nearly one-fifth of GDP.^45^ Many Guatemalans therefore age with adult children living outside the country, household income that depends on remittances, and traditional caregiving arrangements disrupted by familial absences. The impact of these migration dynamics on social care, connectedness, and healthy aging is incompletely understood.

### Study objective

The long-term objective of our research program is to develop ELEGUA into a nationally representative, population-based, longitudinal aging study modeled on the HRS International Network of Studies, including an HCAP subsample. The pilot study described in this protocol advances this objective by testing study procedures, including sampling, survey technology implementation, and data collection in Spanish and Mayan. Pilot findings will inform adaptations to study instruments and protocols and establish acceptability in preparation for the national study.

## METHODS AND ANALYSIS

### Study design

The ELEGUA pilot is a cross-sectional household survey of middle-aged and older adults in Tecpán, Guatemala, conducted from June through December 2026. This protocol was guided by the Strengthening the Reporting of Observational Studies in Epidemiology (STROBE) reporting statement.^46^

### Setting

The pilot was conducted in Tecpán, a municipality of approximately 92,000 people in the Central Highlands department of Chimaltenango. We selected this municipality as the pilot site for several reasons. First, the language composition of the Tecpán population supports a central pilot objective of testing survey procedures, including cognitive assessments, in both Spanish and a Mayan language. Specifically, about half of Tecpán’s adult population speaks Kaqchikel, a Mayan language with over 400,000 speakers nationwide.^47^ Second, Tecpán’s population is diverse with respect to sociodemographic characteristics that are salient in Guatemala, including rural versus urban residence, Maya versus Ladino ethnicity, and educational attainment. This diversity allowed us to pilot study procedures across a broad range of participants. Third, international migration to the United States and Canada is common among adults in Tecpán;^48^ many residents have migrated and later returned, while others remain abroad and send remittances to family members living in the municipality. This context permitted the ELEGUA migration module to be piloted in a population with substantial experience of international migration and receipt of remittances. Fourth, residents in Tecpán have experienced secular transitions in occupation, lifestyle, and diet that have driven increases in cardiometabolic diseases such as obesity, diabetes, and hypertension.^49,50^ The pilot study was therefore able to test health questionnaire items, physical measurements, and blood collection processes in a population with a high underlying burden of these conditions. Fifth, the pilot built on an established collaboration with a local research organization in Tecpán that has expertise conducting household surveys in Mayan languages.^51,52^

### Survey contents

We first produced complete versions of core HRS and HCAP instruments adapted to Guatemalan Spanish and Kaqchikel. We used the questionnaire recommended by the Gateway to Global Aging Data, a resource for harmonizing data from international aging studies, as the foundation for our core instruments.^53^ The questionnaire from the Mexican Cognitive Aging Ancillary Study (Mex-Cog) served as the foundation for our HCAP instrument.^7^ We also drew on additional items and modules from other HRS and HCAP studies.^2,4,6,54–56^ From these complete instruments, we selected modules for the pilot survey within available resources (**Table 1**), prioritizing those requiring cultural and linguistic adaptation^57^ and also retaining the full HCAP cognitive battery to permit psychometric analysis (**Figure 1**). We also included modules for household respondents, physical measurements, and venous blood collection to allow our team to test field procedures and to assess response rates under conditions approximating those of the planned national study.

**Figure 1:** Administration of the matchstick copy test from the ELEGUA cognitive battery. This figure was removed from the preprint per medRxiv policy on photographs of individuals. It is is available from the corresponding author upon request. [Figure1.jpeg] An interviewer administers a visuospatial construction task from the ELEGUA cognitive battery, in which the respondent reproduces a geometric figure using matchsticks. The matchstick copy test was adapted from HCAP studies in South Africa and Nepal,^57,73–75^ as prepilot work indicated that older Guatemalan adults were often unfamiliar or uncomfortable with pencil-and-paper drawing tasks. The photograph is used with signed consent from the participant.

**Table 1:** Survey contents.

| **Respondent** | **Content** |
| --- | --- |
| Individual respondents* | <u>Questionnaire</u>: Demographics, health and healthcare, functional limitations (activities of daily living [ADLs] and instrumental activities of daily living [IADLs]), mental health, health behaviors (smoking, drinking), employment and retirement, discrimination, and cognition (HCAP battery). <u>Physical measurements</u>: Blood pressure, height, and weight. <u>Blood collection</u>: Venous blood for analysis of hemoglobin A1c. |
| Household respondents | <u>Questionnaire</u>: Household roster, primary residence and real estate, household assets, housing characteristics, general income, food consumption, and healthcare expenditures. |
| Informant<sup>†</sup> | <u>Questionnaire</u>: Demographics, Informant Questionnaire on Cognitive Decline in the Elderly (IQCODE), and Blessed Dementia Rating Scale (BDRS). |

\*If the selected individual respondent lacked capacity to provide informed consent, a proxy respondent could complete designated sections of the individual questionnaire on their behalf; these designated sections included modules on demographics, health, health behaviors, employment, and retirement. The HCAP cognitive battery and other sections requiring direct respondent assessment were not administered by proxy.
<sup>†</sup>Informants were nominated by the individual respondent as persons familiar with their health and cognitive functioning, typically a spouse, adult child, or caregiver.

### Eligibility criteria

The pilot survey included respondents in three roles: (1) individual respondents, who completed the main questionnaire, the HCAP cognitive battery, physical measurements, and venous blood collection; (2) household respondents, who responded to questions on household composition, assets, income, and expenditures; and (3) informants, who reported on the individual respondent’s cognitive and daily functioning (**Table 1**). Each household included exactly one individual respondent, selected probabilistically as described below under Sampling design.

Each household could include up to three household respondents, as different modules of the household questionnaire (family structure, housing and consumption, and household finances) could be completed by a different adult most knowledgeable about that topic. Each individual respondent had one informant, who was nominated by the respondent and completed the informant interview. Eligibility criteria were defined separately for each role, and within a household the same adult could fill more than one role. For example, an individual respondent could also serve as a household respondent. Key terms are defined in **Table 2**.

**Table 2:** Definitions of key terms relating to eligibility criteria.

| **Term** | **Definition** |
| --- | --- |
| Household | All persons who usually live in the same dwelling and share food or other essentials (i.e., take meals from a common kitchen), even if work schedules sometimes prevent them from eating together. In contrast, a dwelling (*vivienda*) is the physical housing unit, which may contain one or more separate households. |
| Household resident | A person who has lived in the household for at least 6 months in the last year or intends to live in the household for at least 6 months in the next year. |
| Institutional setting | A place where residents live under an organized arrangement, including settings such as a nursing or long-term care home (*hogar de ancianos/centro para adultos mayores*); a psychiatric hospital or long-stay psychiatric unit (*hospital psiquiátrico/unidad de larga estancia*); a convent, monastery, or other religious residence (*convento/monasterio/residencia religiosa*); or a place of incarceration such as a prison, jail, or detention center (*cárcel/centro de detención*). |
| Proxy | A person who responds on behalf of the individual respondent when the individual respondent lacks capacity to consent to participate in the interview because of physical symptoms (e.g., hearing problems, speech problems, severe illness) or cognitive impairment. When multiple eligible individuals are present to serve as the proxy, preference will go to whoever provides the most regular care or is identified by the family as the primary decision-maker. |
| Spouse | A person who is legally married to (*casado/a*) or in a consensual union (*unido/a*) with the individual respondent. |

#### Individual respondents

The individual respondents were community-dwelling individuals aged 40 years or older who were selected probabilistically in the survey. Specific eligibility criteria were:

1. Household resident
2. Aged 40 years or older
3. Did not live in an institutional setting
4. Able to complete interviews in Spanish or Kaqchikel
5. Provided informed consent, or, if lacking capacity to consent, provided verbal assent with signed consent from a proxy

If the selected individual respondent lacked capacity to provide informed consent, a proxy respondent could complete designated sections of the questionnaire on the individual respondent’s behalf (with respondent’s assent), consistent with procedures used in other HCAP studies.^58^ Proxies were required to be at least 18 years of age, have regular contact with the participant, be familiar with the participant’s health status, family circumstances, and finances, and themselves have capacity to provide consent.

The lower eligibility age bound varies across core studies in the HRS International Network (e.g., 40 years in South Africa,^59^ 45 years in India^55^ and China,^60^ 50 years in Mexico,^6^ and 51 years in the U.S. HRS).^2^ We selected a lower bound of 40 years, matching that used in South Africa, for three primary reasons. First, a central rationale for developing ELEGUA is that early-life nutritional deprivation, widespread poverty, and minimal social protection may accelerate aging processes in Guatemala relative to other settings. Second, cardiometabolic diseases and their impact on aging outcomes are important to characterize, and the prevalence of these conditions rises sharply around age 40 years in Guatemala.^50,61^ Third, retirement may occur at younger ages in Guatemala than in other settings because pensions for civil servants require 20 years of service with no minimum age requirement. Entry into public service around age 20 was common until 2012, particularly among teachers, so many civil servants become eligible for a pension around age 40.^62^

#### Household respondents

The household respondents were adults living in each household who were selected as the most knowledgeable about each household questionnaire module. Specific eligibility criteria were:

1. Household resident
2. Aged 18 years or older
3. Knowledgeable about household members, family structure, and/or finances
4. Able to complete interviews in Spanish or Kaqchikel
5. Provided informed consent

#### Informants

The informant, who was nominated by the individual respondent, provided complementary information on the individual respondent’s cognitive and daily functioning. Specific eligibility criteria were:

1. Aged 18 years or older
2. Nominated by the individual respondent
3. Able to complete interviews in Spanish or Kaqchikel
4. Provided informed consent

### Sampling and sample size

#### Sampling design

The pilot used a multistage stratified cluster sampling design. The sampling frame consisted of the 355 primary sampling units (PSUs) defined by Guatemala’s National Institute of Statistics (INE) in Tecpán. There were an average of 160 dwellings per PSU. The INE provided maps and household listings used for stratification, sample selection, and fieldwork planning. The design was explicitly stratified by urban and rural residence. In addition, the PSUs were sorted geographically before selection, inducing implicit stratification. Every eligible adult in the target population had a non-zero probability of selection.

In the first stage, we selected 30 PSUs by systematic sampling with probability proportional to size, where size is the number of private households recorded for each PSU. Twenty PSUs constituted the main sample; the remaining 10 were held in random order as a supplementary sample that would be released if time and resources permitted. In the second stage, we selected eligible households within each PSU using inverse sampling.^63,64^ Specifically, households enumerated from INE maps were placed in random order within each PSU, and field teams approached them in this order until the target number of 20 eligible and participating households was achieved. In the third stage, we randomly selected one respondent per household using a 2:1 oversampling factor for adults aged 55 years and older.

#### Sample size and precision

While the objective of the pilot survey was to test procedures rather than to achieve a given statistical precision, we designed the sample to also yield informative population estimates that could be shared with local stakeholders. Our calculations conservatively assumed 400 respondents in 20 PSUs. For an illustrative condition with 25% prevalence, such as hypertension, and an intra-cluster correlation of 0.01, the combined design effect from clustering and oversampling weights was estimated to be 1.27, yielding an effective sample size of approximately 315 and a 95% margin of error of ±4.8 percentage points (95% CI 20.2% to 29.8%).

This sample size also supported a preliminary assessment of measurement differences by interview language in the HCAP cognitive battery, specifically, whether item-response probabilities differ between Spanish and Kaqchikel administration, conditional on underlying cognitive ability.^56^ We anticipated interviews to be distributed relatively evenly between the two languages; however, even under a conservative assumption of an uneven distribution (e.g., one-third versus two-thirds), 400 respondents provided more than 80% power (α=0.05, two-sided) to detect differential item functioning of the cognitive tests, corresponding to an odds ratio of 2.0 in a logistic regression framework.

### Data collection

Pilot fieldwork was carried out by a project coordinator, field supervisor, project assistant, driver, and three two-person field teams, each consisting of an interviewer and a nurse. Staff were recruited based on educational qualifications, prior field experience, and Spanish-Kaqchikel bilingualism (including literacy in both languages). Interviewers and nurses completed a two-week training workshop that concluded with a competency assessment, including a supervisor-observed visit to certify each trainee’s ability to administer the HCAP battery.

Questionnaire data were captured on Android tablets using the NubiS computer-assisted personal interviewing platform.^65^ NubiS provides integrated sample management, multilingual electronic data capture, and mapping tools that allow field staff to locate the position of selected households using maps of each primary sampling unit. The platform has been used in HRS and HCAP studies in India, South Africa, China, and many other countries. Data were captured offline, encrypted on the device, and uploaded to a central server when an internet connection became available.

At the start of each visit, interviewers asked respondents to choose the language of administration (Spanish or Kaqchikel), emphasizing that each option is acceptable and that respondents may switch between languages at any time during the interview. A household respondent was interviewed first to complete the household roster and enable random selection of the individual respondent. When feasible, the informant interview was conducted concurrently with the individual interview to improve fieldwork efficiency, reduce interruptions, and preserve the independence of the informant report by ensuring the two respondents were not in the same room. Individual respondents were always interviewed in person to standardize administration of the cognitive battery and permit physical measurements and blood collection; household and informant interviews could be completed by telephone. Before beginning the cognitive module, which included vision-dependent assessments, interviewers asked whether the individual needed glasses to see up close. Those with uncorrected near-vision difficulty were offered free reading glasses to limit potential measurement error attributable to presbyopia. With respondent permission, the cognitive battery and selected gate and quality-control items were audio recorded to permit independent rescoring and monitoring of interviewer performance.

Physical measurements and venous blood were obtained from individual respondents using protocols adapted from other HRS and HCAP studies and INCAP’s protocols previously used in Guatemala.^66,67^ Height was measured using portable stadiometers and weight using digital scales. Blood pressure was measured in triplicate using a validated automated device (HEM-907XL, Omron) with appropriate cuff size. Participants were seated with back supported, feet flat on the floor, and the left arm supported at the level of their heart. Measurements were taken at 1-minute intervals after at least 5 minutes of rest. Nurses trained in phlebotomy collected approximately 2 mL of venous blood by antecubital venipuncture. Blood specimens were transported under cold chain to the INCAP laboratory in Guatemala City and analyzed for hemoglobin A1c on the Roche Cobas c111 system. As the purpose of venous blood collection primarily was to test ELEGUA procedures, no further assays were performed and samples were not stored for future use. Respondents received a printed summary of their measurements and hemoglobin A1c result; those with values in the prediabetes or diabetes range received a call from a physician or study nurse and a written referral to a local primary care facility.

Respondents received supermarket gift cards valued at GTQ 50 (approximately USD $6.50) after completing the questionnaire and physical measurements, and a second card of equal value after venous-blood collection. The amount was chosen to align with the local minimum wage of approximately GTQ 125 per day (approximately USD $16).^68^

### Planned statistical analysis

Population estimates will use sampling weights defined by the selection probabilities at each stage, age-based oversampling, and an adjustment for household eligibility under inverse sampling. We will calibrate weights based on INE population projections derived from the 2018 census. Variance estimation will account for the complex design using Taylor linearization. We will describe feasibility indicators such as household and individual response rates, classified according to AAPOR definitions;^69^ interview duration; item-level missingness; and other indicators. Psychometric analyses of the cognitive battery will proceed in three steps. We will first review item-level response distributions with the field team to identify potential administration problems. We will then fit confirmatory factor analysis models to assess dimensionality and model fit.^70,71^ Finally, we will assess differential item functioning by language of administration, as described above.^56,72^

### Patient and public involvement

Patients and the public were not involved in the development of the protocol. Their involvement in the dissemination of findings is detailed in the Dissemination plan.

## ETHICS AND DISSEMINATION

### Research ethics approval

Ethics approval was obtained from the INCAP Research Ethics Committee, the Maya Health Alliance IRB, and the University of Michigan IRB.

### Informed consent

Many Guatemalans, particularly older adults and Indigenous Maya, have a deeply rooted cultural distrust of signing documents. This distrust stems from historical experiences of exploitation, coercion, and dispossession related to land and other resources, experiences which were especially marked during the Guatemalan Civil War. This context informed our tiered consent approach: Verbal consent was used for questionnaire assessments and signed consent was required only for blood collection or use of a proxy respondent. When a proxy was used for a respondent who lacked capacity, the proxy respondent provided signed consent and the respondent provided verbal assent. Bilingual field staff always administered consent in the respondent’s preferred language (Spanish or Kaqchikel).

### Data monitoring and safety

The ELEGUA pilot survey was a minimal-risk observational study, and adverse events were not expected. Nonetheless, the study team documented any adverse events on a reporting form and followed the reporting guidelines of each supervising IRB. A general physician licensed in Guatemala provided clinical monitoring and, when appropriate, referrals. The investigator team also tracked protocol deviations, complaints, and other reportable information and reported these to the IRBs.

### Data sharing and dissemination plan

We plan to share de-identified data and accompanying documentation from the ELEGUA pilot survey in a public repository, such as the Gateway to Global Aging or Harvard Dataverse.

Results will be published in peer-reviewed journals following established reporting guidelines and presented at national and international conferences. Within Guatemala, we will present findings to the Ministry of Health, the National Committee for the Protection of the Elderly, and other key stakeholders. Regionally in Latin America, we will share results through public-sector technical networks in which INCAP participates, such as the Technical Commission on Chronic Diseases and Cancer, which is composed of delegates from the ministries of health and social security institutes of Central American countries. Locally in Tecpán, we will hold public meetings with municipal and village leaders to discuss findings and their implications.

## ADDITIONAL INFORMATION

### Contributors

DF, EB, MRZ, and JB wrote the funding proposals. DF and KCorzantes wrote the first draft of the pilot survey protocol. KChoy, SA, LFC, AG, KL, PR, and BW contributed to the research design and the development of study procedures. All authors reviewed multiple iterations of the research application and protocol, and all approved the final manuscript.

### Patient consent for publication

The respondent shown in Figure 1 provided signed consent for the use of this image.

## Supporting information

STROBE checklist

