## Supplementary material for "Developing the Longitudinal Study of Aging in Guatemala (ELEGUA): Rationale and pilot protocol": STROBE checklist

STROBE Statement—Checklist of items that should be included in reports of ***cross-sectional studies***

*Manuscript: Developing the Longitudinal Study of Aging in Guatemala (ELEGUA): Rationale and pilot protocol. Locations are given as manuscript section/subsection titles rather than page numbers so they remain valid under repagination. Results-stage items are marked N/A because this is a protocol paper.*

|  | Item No | Recommendation | Section |
| --- | --- | --- | --- |
| **Title and abstract** | 1 | (*a*) Indicate the study’s design with a commonly used term in the title or the abstract | Title (“pilot protocol”); Abstract – Methods and analysis (“cross-sectional household survey”) |
|  |  | (*b*) Provide in the abstract an informative and balanced summary of what was done and what was found | Abstract |
| Introduction | | | |
| Background/rationale | 2 | Explain the scientific background and rationale for the investigation being reported | Introduction; Introduction – Rationale for a longitudinal aging study in Guatemala |
| Objectives | 3 | State specific objectives, including any prespecified hypotheses | Abstract – Introduction; Introduction – Study objective |
| Methods | | | |
| Study design | 4 | Present key elements of study design early in the paper | Methods and analysis – Study design |
| Setting | 5 | Describe the setting, locations, and relevant dates, including periods of recruitment, exposure, follow-up, and data collection | Methods and analysis – Study design; Methods and analysis – Setting |
| Participants | 6 | (*a*) Give the eligibility criteria, and the sources and methods of selection of participants | Methods and analysis – Eligibility criteria; Methods and analysis – Sampling design; Table 2 |
| Variables | 7 | Clearly define all outcomes, exposures, predictors, potential confounders, and effect modifiers. Give diagnostic criteria, if applicable | Methods and analysis – Survey contents; Table 1 |
| Data sources/ measurement | 8* | For each variable of interest, give sources of data and details of methods of assessment (measurement). Describe comparability of assessment methods if there is more than one group | Methods and analysis – Survey contents; Methods and analysis – Data collection |
| Bias | 9 | Describe any efforts to address potential sources of bias | Methods and analysis – Data collection; Methods and analysis – Planned statistical analysis |
| Study size | 10 | Explain how the study size was arrived at | Methods and analysis – Sample size and precision |
| Quantitative variables | 11 | Explain how quantitative variables were handled in the analyses. If applicable, describe which groupings were chosen and why | Methods and analysis – Planned statistical analysis; Methods and analysis – Sampling design |
| Statistical methods | 12 | (*a*) Describe all statistical methods, including those used to control for confounding | Methods and analysis – Planned statistical analysis |
|  |  | (*b*) Describe any methods used to examine subgroups and interactions | Methods and analysis – Planned statistical analysis |
|  |  | (*c*) Explain how missing data were addressed | Methods and analysis – Planned statistical analysis |
|  |  | (*d*) If applicable, describe analytical methods taking account of sampling strategy | Methods and analysis – Planned statistical analysis |
|  |  | (*e*) Describe any sensitivity analyses | N/A – no sensitivity analyses prespecified in this pilot protocol |
| Results | | | |
| Participants | 13* | (a) Report numbers of individuals at each stage of study—eg numbers potentially eligible, examined for eligibility, confirmed eligible, included in the study, completing follow-up, and analysed | N/A – protocol paper |
|  |  | (b) Give reasons for non-participation at each stage | N/A – protocol paper |
|  |  | (c) Consider use of a flow diagram | N/A – protocol paper |
| Descriptive data | 14* | (a) Give characteristics of study participants (eg demographic, clinical, social) and information on exposures and potential confounders | N/A – protocol paper |
|  |  | (b) Indicate number of participants with missing data for each variable of interest | N/A – protocol paper |
| Outcome data | 15* | Report numbers of outcome events or summary measures | N/A – protocol paper |
| Main results | 16 | (*a*) Give unadjusted estimates and, if applicable, confounder-adjusted estimates and their precision (eg, 95% confidence interval). Make clear which confounders were adjusted for and why they were included | N/A – protocol paper |
|  |  | (*b*) Report category boundaries when continuous variables were categorized | N/A – protocol paper |
|  |  | (*c*) If relevant, consider translating estimates of relative risk into absolute risk for a meaningful time period | N/A – protocol paper |
| Other analyses | 17 | Report other analyses done—eg analyses of subgroups and interactions, and sensitivity analyses | N/A – protocol paper |
| Discussion | | | |
| Key results | 18 | Summarise key results with reference to study objectives | N/A – protocol paper |
| Limitations | 19 | Discuss limitations of the study, taking into account sources of potential bias or imprecision. Discuss both direction and magnitude of any potential bias | Strengths and limitations of this study |
| Interpretation | 20 | Give a cautious overall interpretation of results considering objectives, limitations, multiplicity of analyses, results from similar studies, and other relevant evidence | N/A – protocol paper |
| Generalisability | 21 | Discuss the generalisability (external validity) of the study results | Strengths and limitations of this study; Methods and analysis – Sample size and precision |
| Other information | | | |
| Funding | 22 | Give the source of funding and the role of the funders for the present study and, if applicable, for the original study on which the present article is based | Additional information – Funding |

*Give information separately for exposed and unexposed groups.

**Note:** An Explanation and Elaboration article discusses each checklist item and gives methodological background and published examples of transparent reporting. The STROBE checklist is best used in conjunction with this article (freely available on the Web sites of PLoS Medicine at http://www.plosmedicine.org/, Annals of Internal Medicine at http://www.annals.org/, and Epidemiology at http://www.epidem.com/). Information on the STROBE Initiative is available at www.strobe-statement.org.
